# Prevalence, risk factors, and impact of anxiety in early Alzheimer disease: a retrospective study of an autopsy-confirmed cohort

**DOI:** 10.1101/2024.08.04.24311473

**Authors:** Palak Patel, Mark A. Bernard, Arjun V. Masurkar

**Affiliations:** Alzheimer’s Disease Research Center, NYU Grossman School of Medicine, New York, NY, 10016; Center for Cognitive Neurology, Department of Neurology, NYU Grossman School of Medicine, New York, NY, 10016; Neuroscience Institute, NYU Grossman School of Medicine, New York, NY, 10016; Departments of Neurology and Psychiatry, Hackensack Meridian School of Medicine, Nutley, NJ 07110

**Keywords:** Alzheimer disease, neuropathology, anxiety, subjective cognitive decline, mild cognitive impairment, dementia

## Abstract

Anxiety is a neuropsychiatric symptom (NPS) of Alzheimer disease (AD) patients which has been studied primarily in prospective and retrospective studies of clinically diagnosed AD. However, this can be confounded by other primary etiologies. Moreover, anxiety has not been comprehensively studied in autopsy-confirmed AD cases across subjective cognitive decline (SCD), mild cognitive impairment (MCI), and dementia stages. We conducted a retrospective longitudinal analysis of 212 participants with autopsy-confirmed AD, followed from 1986-2013 at the NYU Alzheimer’s Disease Research Center with staging via the Global Deterioration Scale and NPS assessed via BEHAVE-AD. We found that anxiety varied uniquely with stage and was the most common NPS in SCD and MCI (35-40% prevalence). ApoE4 carriage associated with a higher rate of anxiety only at mild dementia. Anxiety in SCD associated with cerebral amyloid angiopathy and arteriosclerosis on brain autopsy, but there were no such associations with concomitant neuropathology at MCI and mild dementia. Anxiety associated with increased progression rate (∼2.5-fold) from SCD to MCI/dementia stages, but not from MCI to dementia. These results suggest an important relationship between anxiety and AD, especially at the preclinical stage. This warrants further study of anxiety as a possible modifiable factor of disease experience and course.

## Introduction

Neuropsychiatric symptoms (NPS) are well-known features of Alzheimer disease (AD) (Lyketsos et al., 2002). Studies of NPS in clinically diagnosed AD have led to several observations. These symptoms follow a contrasting trajectory to cognition and function by progressively increasing and peaking before the terminal stages of AD (Reisberg et al., 2014). Certain symptoms such as depression and anxiety predominate in early stages while psychosis and agitation are common in later stages (Selbaek et al., 2014). Depression and anxiety symptoms are additionally observed in cognitively normal elderly population (Geda et al., 2008). Given their temporal evolution, NPS have been investigated as risk factors of cognitive decline in a number of studies (Geda et al., 2008; Burhanullah et al., 2019). Depression in particular has been the focus of a number of studies, but evidence to support it as a risk factor has been inconsistent (Devanand et al., 1996; Modrego and Ferrandez 2004; Green et al., 2003; Teng et al., 2007; Wilson et al., 2008; Panza et al., 2008; Rozzini et al., 2005; Becker et al., 2009).

Despite being frequently comorbid with depression in AD (Ferretti et al., 2001), the role of anxiety initially was less studied (Cohen 1998). Anxiety has since been observed to be correlated with progression to clinically-defined AD from preclinical stages (Donovan et al., 2014; Mah et al., 2015; Santabarbara et al., 2019) and has associations with AD biomarkers (Patel and Masurkar, 2021). Yet, it remains unclear if anxiety is an early feature of AD (Gallagher et al., 2011), shares common risk factors for dementia such as ApoeE and cerebrovascular disease (Michels et al., 2012, Johansson et al., 2019), or rather is a reaction to cognitive decline (Palmer et al., 2007). Furthermore, the prevalence (Apostolova and Cummings, 2008) and nature of anxiety are also variable across studies examining clinically diagnosed cohorts due to differences in methodologies and population studied (Palmer et al., 2007, Gallagher et al., 2011; Snitz et al., 2018) or the occurrence of other dementia pathologies alone or in conjunction with AD (Becker et al., 2019; Segers et al., 2020). As a definitive diagnosis of AD can only be made by neuropathologically (Jack et al., 2018), a systematic evaluation of anxiety in pathologically proven AD may help to clarify these discrepancies and inform uniformity. We thus sought to leverage an autopsy-confirmed AD cohort to study the distribution of anxiety versus other NPS across the dementia spectrum, characterize the specific anxiety symptoms found in early stages of AD, examine its relation to demographic and genetic factors, assess the effect of secondary pathologies, and quantify the relationship of anxiety symptoms to AD progression from early stages.

## Material and Methods

The study was performed under an approved protocol for the NYU Alzheimer’s Disease Research Center (ADRC) from the Institution Review Board of the NYU Grossman School of Medicine.

### Subjects

We reviewed retrospective longitudinal data on 212 subjects with autopsy-confirmed AD who were enrolled in the NYU ADRC study and followed between 1986 to 2013 by the Clinical Core. These subjects had been recruited from the community via advertisement or referral. During this period, inclusion and exclusion criteria to the study were as follows. Inclusion criteria were age > 40, availability of study partner, fluency of the subject and study partner in English or Spanish, and willingness to undergo phlebotomy and neuroimaging (CT head or MRI brain). Exclusion criteria were history of significant traumatic brain injury, epilepsy, mental retardation, presence of other significant neurological disease, modified Hachinski Ischemia Score ≥4 or history of significant stroke, significant drug or alcohol abuse, significant psychiatric disease, significant medical problems or medications that could affect cognition. Once enrolled, visits included clinical assessment, psychometric testing, and apolipoprotein E (APOE) genotyping. Autopsy was performed by the ADRC Neuropathology Core upon brain donation.

### AD staging and neuropsychiatric assessments

Per NYU ADRC protocol these subjects were staged using the Global Deterioration Scale (GDS) (Reisberg, 1982). GDS consists of 6 stages with GDS 1 correlating to normal cognition, GDS 2 to subjective cognitive decline (SCD), GDS 3 to mild cognitive impairment (MCI), GDS 4 to mild dementia, GDS 5 to moderate dementia, GDS 6 to moderately severe dementia and GDS 7 to severe dementia. Consensus diagnosis along the GDS spectrum was based on clinical evaluation and psychometric testing. Neuropsychiatric symptoms assessments were conducted using the Behavioral Pathology in Alzheimer’s Disease (BEHAVE-AD) instrument (Reisberg et al., 1987). It is a comprehensive, informant-based scale that identifies and assesses the behavioral and psychological symptoms noted over the prior two weeks (Reisberg et al., 1987). It scores 25 symptoms on a 4-point severity scale in the following 7 categories: (A) Paranoid and Delusional Ideation, (B) Hallucinations, (C) Activity Disturbances, (D) Aggressiveness, (E) Diurnal Rhythm Disturbances, (F) Affective Disturbances (Depression), and (G) Anxieties and Phobias. For the purpose of analysis, groups (A) and (B) were combined as Psychosis.

### Statistical analyses

Descriptive analyses were used to compare prevalence of neuropsychiatric symptoms at various GDS stages. Student T-test was used to compare scalar measures between groups. Chi squared was used to compare the prevalence of a dichotomous measure between groups. For progression rate, participants with longitudinal visits were included and two forms of analysis were performed. In one form, stage worsening was considered as a binary event and Kaplan-Meier analysis was applied. In the other, a progression rate was calculated as change in GDS divided by total followup time. The p-value for the relationship between anxiety and mean progression was corrected for covariates via a multivariable linear regression model including anxiety presence, age, sex, education, ApoE4 carrier status. All statistical analyses were performed using Prism version 10 software.

## Results

### Prevalence of anxiety versus other symptoms across AD stages

We examined the prevalence of anxiety and fear (a related symptom) versus other NPS categories across AD stages as defined by the Global Deterioration Scale or GDS. NPS categories were assayed via the BEHAVE-AD scale as delineated in Methods. In Figure 1A, we show the relative prevalence of each NPS category within each GDS stage. The anxiety and fear category were the dominant symptom in stage 2 (subjective cognitive decline or SCD), as well as in stage 3 (mild cognitive impairment or MCI), where it was reported in 40% of participants. It was overtaken by activity disturbance in stage 4 (mild dementia) and also psychosis in stage 6 (moderately severe dementia). Thus, anxiety and fear appear to be a neuropsychiatric hallmark of early stages of AD. In Figure 1B, we show this same data but with axes reversed to demonstrate the evolution of each NPS category across stages. Anxiety and fear, and nearly all other symptom categories, strictly increased in prevalence from the GDS stage 2 to stage 5 (moderate dementia), albeit with different rates. However, along with depression, it was the only category to peak in stage 5. In contrast, all other NPS categories increased through stage 6. The final stage 7 (severe dementia) featured a decline in all NPS categories. In sum, anxiety and fear have a unique temporal profile during the course of AD, being the dominant NPS and only affective symptom in preclinical SCD stage and increasing to peak in the moderate dementia stage.

**Figure 1:**
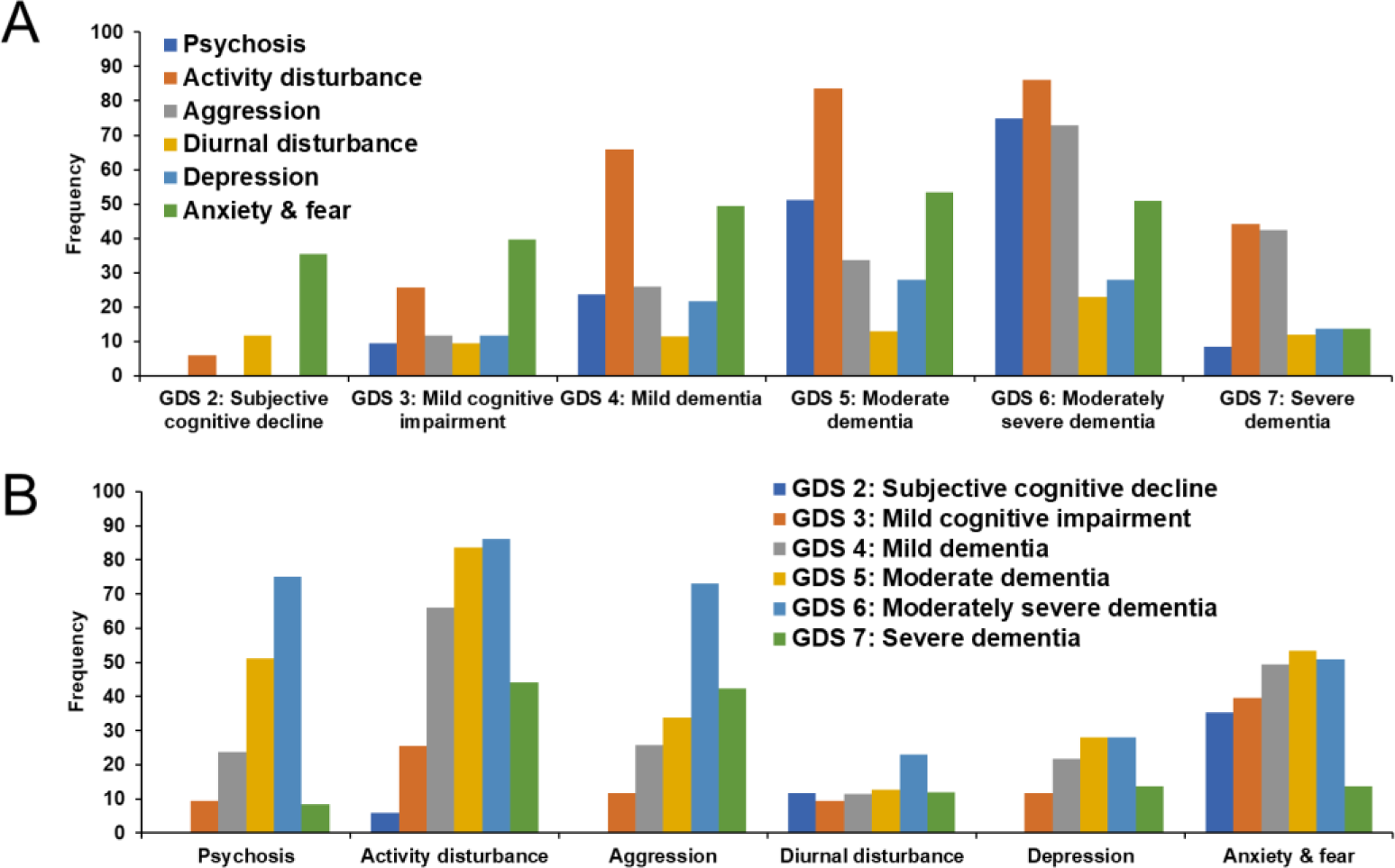
Neuropsychiatric symptoms (NPS) by Global Deterioration Scale (GDS) stage and symptom category. **A.** For each GDS stage, frequency of each NPS category is demonstrated. **B.** For each NPS category, frequency across GDS stage is shown. GDS 2: n=18; GDS 3: n=43; GDS 4: n=97; GDS 5: n=86; GDS 6: n=100; GDS 7: n=59

We also examined individual responses within the anxiety and fear category. The BEHAVE-AD scale differentiates these into four symptoms: Godot syndrome (anxiety for upcoming events), other anxieties (related to money, future, being away from home, health, memory or generalized anxiety such as thinking everything is “terribly wrong”), fear of being alone, and other phobias (crowds, travel, darkness, people/strangers, bathing, etc). We found that “other anxieties” was the sole subcategory in the SCD stage and was the dominant symptom at all stages, whereas Godot syndrome first appeared in MCI and was overall the second most common symptom through GDS stage 6 (Figure 2A). Across stages, the anxiety subcategories declined after stage 5 whereas fears and phobias peaked or plateaued through stage 6 (Figure 2B). These data support a temporal distinction between anxiety and fears, with the former playing a dominant role in early stages.

**Figure 2:**
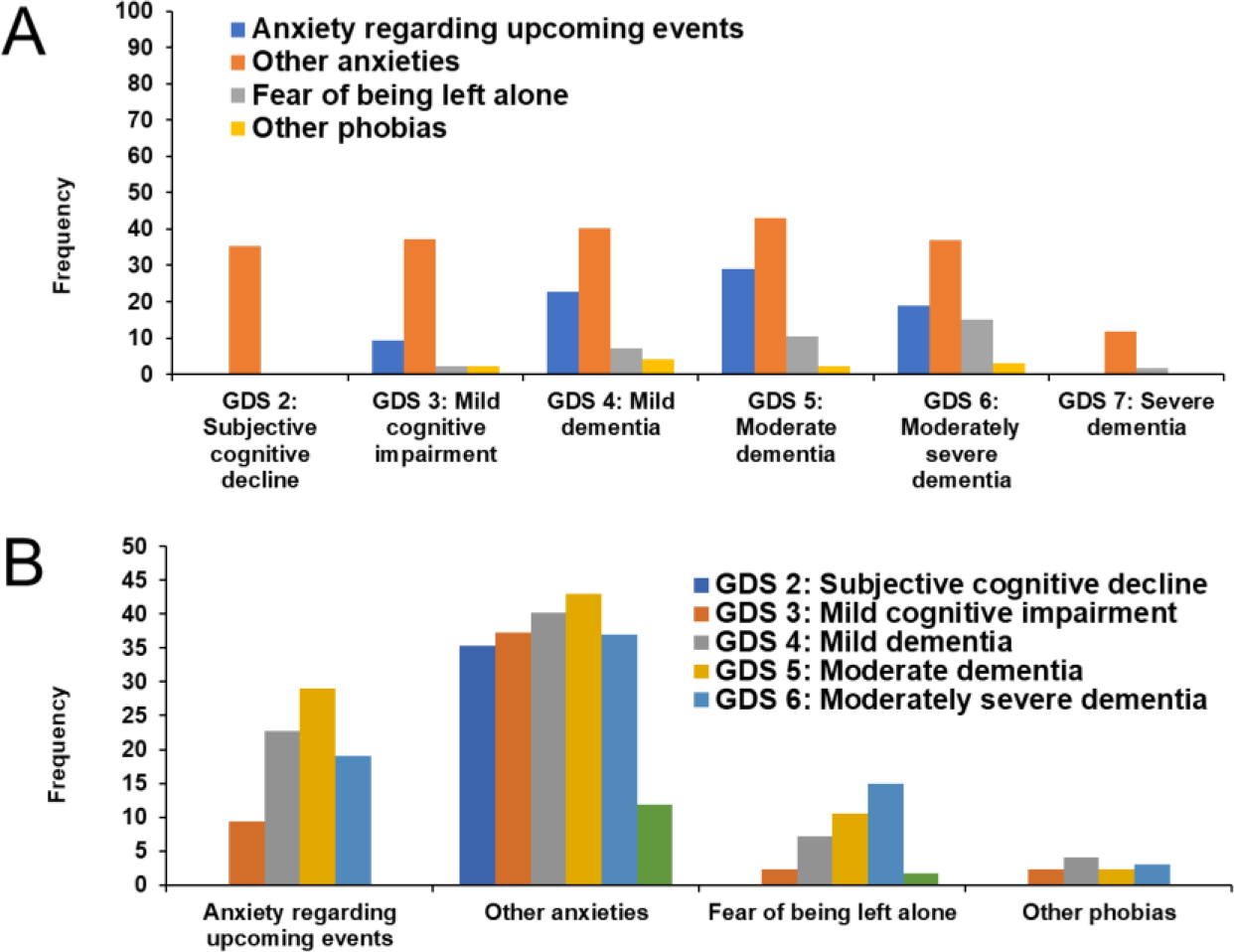
Anxiety and fear symptoms by GDS stage and NPS subcategory. **A.** Across each GDS stage, frequency of each anxiety & fear symptom is displayed. **B.** In the anxiety & fear subcategory, for each specific symptom the frequency across GDS stage is presented. GDS 2: n=18; GDS 3: n=43; GDS 4: n=97; GDS 5: n=86; GDS 6: n=100; GDS 7: n=59

Since anxiety and depression often co-occur, we also analyzed the rate at which symptoms in these two categories presented independently across stages. Consistent across stages (Table 1), anxiety was more likely to occur independent of depressive symptoms, though less so in later stages. In contrast, depression was more likely to occur with anxiety than without it, except at the severe dementia stage.

**Table 1:**
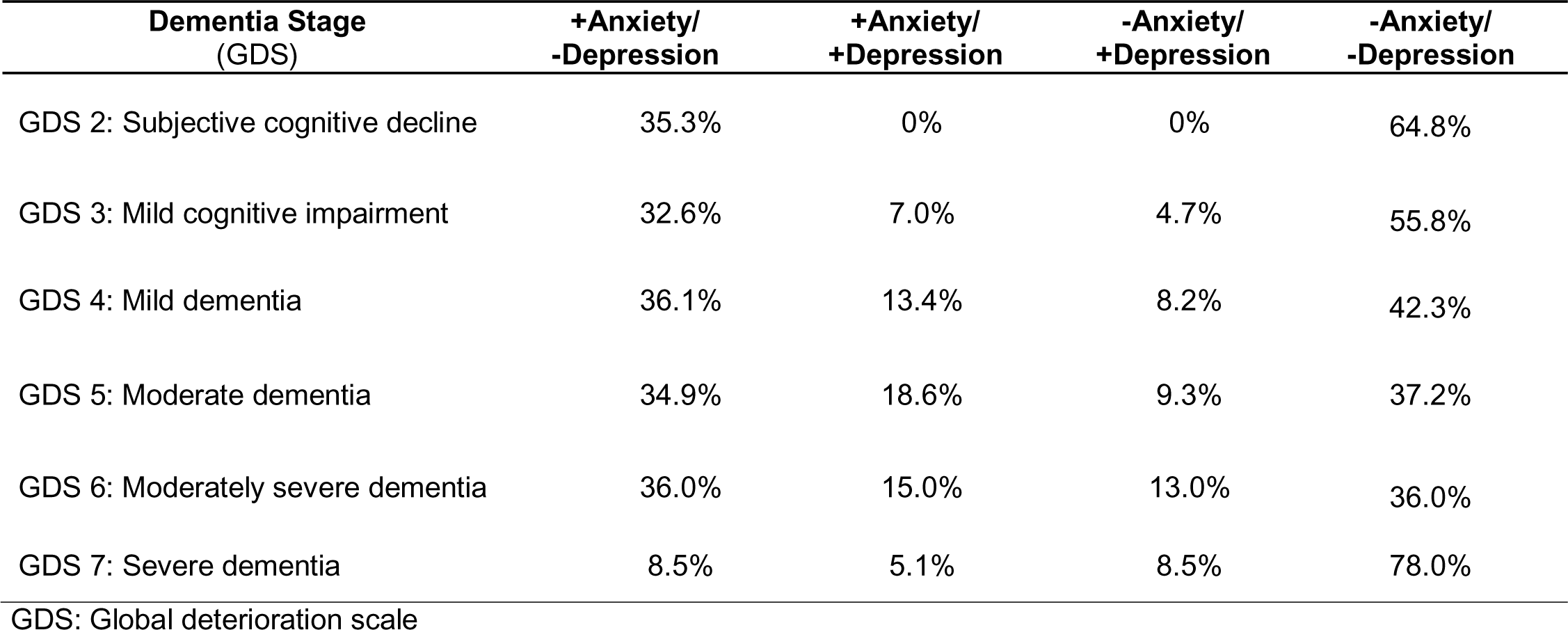
Frequency of anxiety and depression relationships across GDS stage.

### Demographic features associated with anxiety across different stages

We next asked if there were demographic risk factors for anxiety, and if these varied in early versus late stages. These are summarized in Table 2. With regard to age, there were no statistically significant differences between non-anxious and anxious subjects at any stage. Of note, at MCI, anxious subjects were noted to be over 6 years younger than non-anxious subjects, however this just missed statistical significance (p = 0.0504). Sex differences were noted at mild dementia and severe dementia. In mild dementia, females were more likely to display anxiety than males (p = 0.0385). A similar trend was seen in SCD, but this did not reach statistical significance (p = 0.09). In contrast, in severe dementia male subjects were more likely to display anxiety (p = 0.002). Trends in educational differences were noted, with anxiety associated with fewer years of education from SCD through moderate dementia and the reverse in the last two stages. However this only reached statistical significance in SCD (p = 0.049).

**Table 2:** Demographic differences between non-anxious and anxious participants across GDS stage.

| Dementia Stage<br>(GDS) | Age (SEM) |  | % Female |  | Years of Education (SEM) |  |
| --- | --- | --- | --- | --- | --- | --- |
|  | -Anxiety | +Anxiety | -Anxiety | +Anxiety | -Anxiety | +Anxiety |
| GDS 2: Subjective cognitive decline | 74.2 (1.9) | 77.1 (3.7) | 45% | 83% | **16.8 (0.6)** | **14.7 (0.8)** |
| GDS 3: Mild cognitive impairment | 78.4 (2.0) | 72.0 (2.3) | 44% | 38% | 16.1 (0.6) | 15.2 (0.6) |
| GDS 4: Mild dementia | 78.6 (1.4) | 78.0 (1.4) | **48%** | **69%** | 15.4 (0.5) | 14.5 (0.4) |
| GDS 5: Moderate dementia | 74.8 (1.4) | 76.4 (1.5) | 45% | 61% | 14.8 (0.6) | 14.5 (0.5) |
| GDS 6: Moderately severe dementia | 79.7 (1.1) | 77.6 (1.3) | 56% | 67% | 13.1 (0.4) | 14.1 (0.6) |
| GDS 7: Severe dementia | 80.1 (1.2) | 74.5 (3.9) | **73%** | **14%** | 13.6 (0.5) | 15.9 (1.1) |

GDS: Global deterioration scale; SEM: Standard error of the mean. Bold indicates p<0.05.

### Role of ApoE4 and anxiety

Apolipoprotein allele variant E4 (ApoE4), beyond being a risk factor for AD, has also been associated with increased anxiety in clinically defined AD (Robertson et al., 2005; Michels et al., 2012; Holmes et al., 2016). We therefore examined whether ApoE4 had an impact on the prevalence of anxiety in autopsy-confirmed AD at various stages. We limited our analysis to only the “other anxieties” symptom, as it was the most prevalent in this category and nearly all cases with Godot syndrome also had “other anxieties”. This analysis (Figure 3) revealed that while there was a trend for higher anxiety in ApoE4 positive individuals, this reached statistical significance only at GDS stage 4 (p = 0.048). To determine if this was unique to anxiety, we performed a similar analysis within the other neuropsychiatric symptom categories. Such relationships at the mild dementia stage were not seen with depression, psychosis, aggression, activity disturbance, or diurnal disturbance symptom categories (not shown).

**Figure 3:**
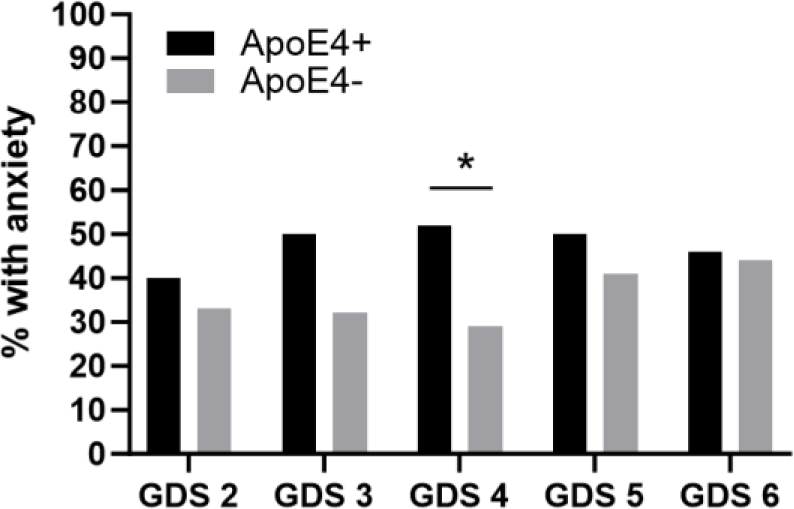
ApoE4 and anxiety across GDS stages. Bar chart showing prevalence of anxiety in APOE+ and APOE4-individuals at each GDS stage. GDS 2: n=18; GDS 3: n=39; GDS 4: n=70; GDS 5: n=57; GDS 6: n=66. *p < 0.05

### Neuropathological relationships

Concomitant pathologies, either related to AD, such as cerebral amyloid angiopathy (CAA), or other disease such as vascular disease and alpha synucleinopathy, can be associated with unique NPS profiles and thus may drive anxiety in AD patients. We examined the association of additional secondary pathologies post-mortem with the retrospective presence of anxiety at the SCD (Figure 4A), MCI (Figure 4B) and mild dementia stages (Figure 4C). Overall, there were a diverse number of additional pathologies. The most common were AD-related cerebral amyloid angiopathy and cerebrovascular arteriosclerosis. Anxiety in SCD was associated with an increased likelihood of CAA and arteriosclerosis at autopsy. However, at MCI and mild dementia stages, no secondary pathology at post-mortem analysis was associated with a higher retrospective likelihood of anxiety.

**Figure 4:**
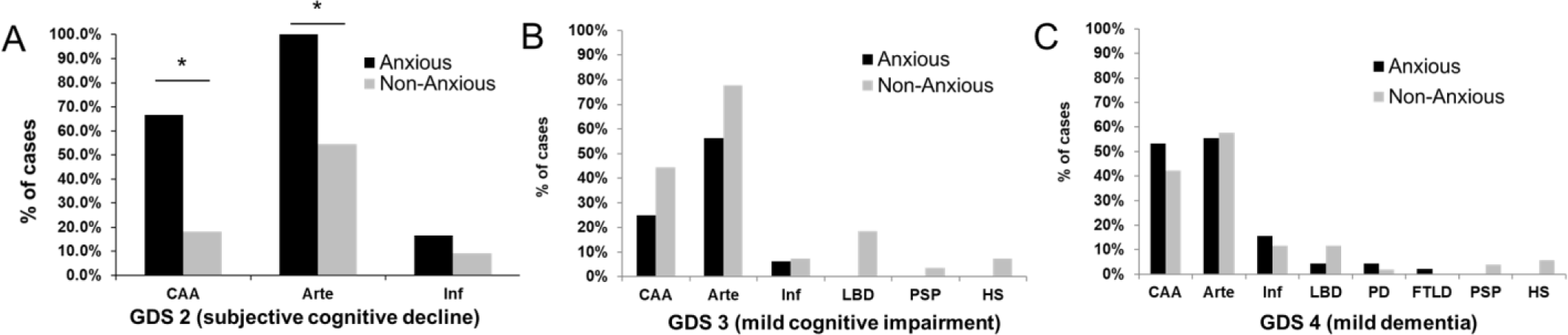
Concomitant neuropathologic diagnoses and likelihood of anxiety in SCD, MCI and mild dementia from AD. **A.** Distribution of concomitant pathology in SCD individuals with and without anxiety. Anxious: n = 6; Non-anxious: n = 11. **A.** Distribution of concomitant pathology in MCI individuals with and without anxiety. Anxious: n = 16; Non-anxious: n = 27. **B.** Distribution of concomitant pathology in mild dementia subjects with and without anxiety. Anxious: n = 45; Non-anxious: n = 52. CAA = cerebral amyloid angiopathy; Arte = arteriosclerosis; Inf = infarcts; LBD = Lewy body disease; PD = Parkinson’s disease; FTLD = frontotemporal lobar degeneration; PSP = progressive supranuclear palsy; HS = hippocampal sclerosis. *p < 0.05

### Anxiety and impact on progression

We next asked whether the presence of anxiety in the preclinical SCD or prodromal MCI stage would correlate with increased progression to dementia. There were 17 SCD subjects (6 with anxiety) who had longitudinal followup. With progression treated as a binary event, the Kaplan-Meier curve (Figure 5A) supported a trend for anxious SCD subjects to progress faster, though it did not reach statistical significance (p = 0.1423). Analysis of progression rate (change in GDS units/year) did reveal a ∼2.5 fold faster progression rate in anxious SCD subjects (Figure 5B, p = 0.0249). In contrast, analysis of 39 MCI subjects (14 with anxiety) who had longitudinal followup did not show any increased association with progression via either method (Figure 5C, p = 0.6875; Figure 5D, p = 0.6870).

**Figure 5:**
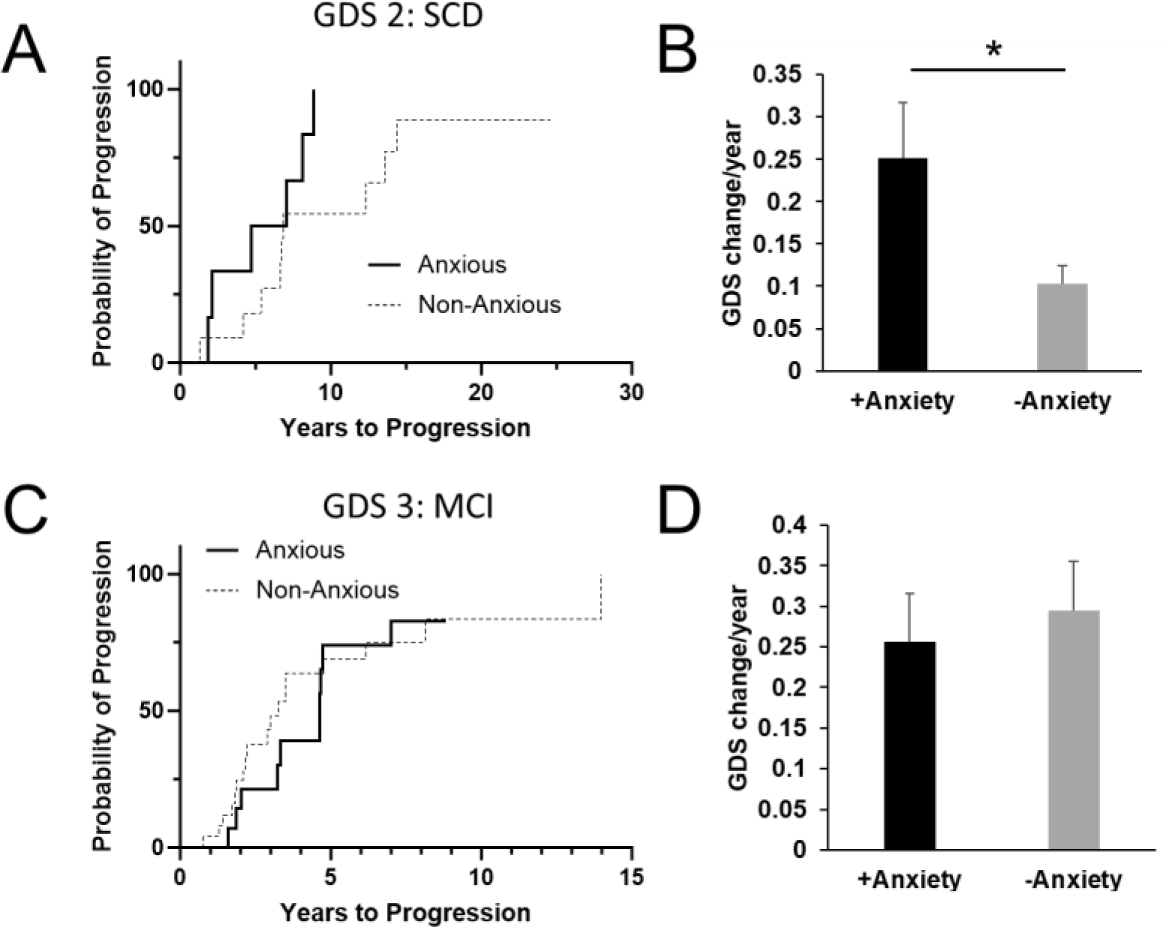
Association of anxiety with progression from SCD and MCI stages. **A**. Kaplan-Meier curve of progression between anxious (n = 6) and non-anxious (n = 11) SCD subjects. **B.** Average progression rate between anxious and non-anxious SCD subjects **C**. Kaplan-Meier curve of progression between anxious (n = 14) and non-anxious (n = 25) MCI subjects. **D.** Average progression rate between anxious and non-anxious MCI subjects. *p<0.05

## Discussion

In this study, we analyzed the prevalence and distribution of anxiety versus other NPS in patients with neuropathologically diagnosed AD, as well as associations of pre-dementia anxiety with ApoE4, neuropathology, and progression rate. Compared to other NPS, anxiety had a unique profile across stages and was the dominant symptom at SCD and MCI, occurring mostly independently of depression. At these pre-dementia stages, demographics and ApoE4 carrier status played a minimal role to influence anxiety prevalence, except that less education correlated with higher anxiety at SCD. Lastly, in contrast to MCI, anxiety at SCD associated with concomitant pathology on autopsy (CAA, arteriosclerosis), and faster progression rate.

Our findings on NPS distribution across AD stages are similar to those in the original retrospective clinical study that led to the development of BEHAVE-AD (Reisberg et al., 1987), as well as in studies of clinical MCI and AD dementia using the Neuropsychiatric Inventory (NPI) as an assessment tool (Lyketsos et al., 2002). In the latter, rates of NPS were lower than our study, including anxiety (5% in healthy, 10% in MCI and 25% in dementia), but consistent with other studies using the NPI (Lyketsos et al., 2000; Seignourel et al., 2008; Geda et al., 2008).

Thus differences may be due to instruments used. Moreover, some studies did not differentiate dementia severity and may have included participants in more severe stages at which anxiety prevalence declines considerably. Consistent across all studies is the early predominance of affective symptoms, with psychosis or agitation emerging later in the course. As will be discussed in subsequent sections, if NPS arise directly from AD pathology itself, this temporal evolution may reflect tauopathic progression from locus coeruleus to medial temporal lobe to neocortex (Braak and Braak 1991; Lopez et al., 2001; Lopez et al., 2005; Ehrenberg et al., 2018).

Our finding that ApoE4 carrier status does not impact anxiety prevalence in SCD and MCI stages differs somewhat from prior studies (Holmes et al., 2016; Michels et al., 2012). Findings may differ due to the use of different scales, lack of autopsy confirmation of diagnoses, and evaluation of cognitively normal but not specifically SCD individuals. In contrast, our finding of ApoE4 carriage increasing anxiety prevalence in the mild dementia stage is consistent with other studies (Robertson et al., 2005; Michels et al., 2012). There may in addition be a cross-interaction with sex, as alluded to in other studies (Robertson et al., 2005), as females also tended to show more anxiety at the mild dementia stage as well. As ApoE4 is associated with mechanisms of early and greater amyloid deposition (Morris et al., 2010; Lim et al., 2017) and network hyperactivity (Nuriel et al., 2017), this may suggest that these processes give rise to the anxiety phenotype in anatomical networks involved in AD specific to the mild dementia stage and that other mechanisms may play a role earlier.

Interestingly, we additionally found that prevalence of anxiety in SCD, but not MCI, was at least partially associated with eventual concomitant pathology, namely CAA and arteriosclerosis, on brain autopsy. Considering the delay between SCD and death, we cannot assume this pathology was present at the SCD stage. However, risk factors for these neuropathologies may co-occur with anxiety at these stages, for example vascular comorbidities. Our findings are nevertheless supported by other studies that observed elevated anxiety in patients with CAA (Smith et al., 2021) and another study that reported elevated likelihood of anxiety in cognitively normal patients with white matter hyperintensities on MRI (Liampas et al., 2024), which may reflect arteriosclerosis or CAA (Charidimou et al., 2016). Of note, evidence that anxiety arises from parenchymal amyloid alone in normal adults is mixed (Pietrzak et al., 2015; Hanseeuw et al., 2020; Johansson et al., 2020; Krell-Roesch et al., 2018; Bensamoun et al., 2016) and has more evidence at the MCI stage Bensamoun et al., 2016; Krell-Roesch et al., 2019; Goukasian et a., 2019; Johansson et al., 2020; Ramakers et al., 2013). This does not rule out that primary AD pathology plays a critical role, as a prior study implicated early stage tauopathy in the development of anxiety (Ehrenberg et al., 2018).

Our finding in this autopsy-confirmed cohort that anxiety associates with faster progression from the SCD stage is consistent with prior studies examining normal/preclinical persons without autopsy confirmation of eventual AD diagnosis (Donovan et al., 2014; Sinoff and Werner, 2003; Snitz et al., 2018; Liew 2020). Of note, one of these studies found that the presence of SCD itself potentiated the impact of anxiety (Liew 2020), and another noted that anxiety about memory (Snitz et al., 2018) was particularly predictive of decline in the SCD stage (Snitz et al., 2018). While we did not assess this specific anxiety in our study, this symptom has been cited as part of the SCD-plus criteria that raise the likelihood that SCD correlates with AD biomarkers (Jessen et al., 2014). In contrast, we find no impact of anxiety on progression from the MCI level. Of note, prior studies without autopsy-confirmed AD outcomes have also had mixed results based on specific anxiety symptoms and time interval (Palmer et al., 2007; Wadsworth et al., 2012; Donovan et al., 2014; Mah et al., 2015; Rosenberg et al., 2013; Somme et al., 2013; Ramakers et al., 2010; Devier et al., 2009). Overall, our results in addition to the above studies strongly suggests that persons with SCD and anxiety are an at-risk group warranting further study, and specifically at the level of mechanism and biomarker evolution.

Our study has multiple strengths and a few limitations. Strengths include a robustly characterized clinically cohort, identification of the SCD stage, broad phenotyping of NPS, autopsy-based diagnoses, inclusion of ApoE genotype, and robust sample size for dementia stages. Limitations include smaller sample size for early stages, biases on which participants may come to autopsy, NPS reporting based on informant as opposed to participant report, and lack of cross-sectional data on regional distribution of AD pathology (amyloid or tau PET), which was not widely available during the period of the longitudinal study.

Overall, these results suggest an important relationship between anxiety and AD, especially at preclinical and prodromal stages. As such, anxiety may be an important clinical marker of a more aggressive form of AD, or perhaps is direct influencer of disease progression. This warrants further study, as improved treatments of anxiety in early stages may not only improve patient quality of life but could also have important disease modifying capabilities.

## Data Availability

All data produced in the present study are available upon reasonable request to the authors

